# Botulinum Toxin Application for Treatment of Graft Vasospasm: A Reverse Translational Study

**DOI:** 10.1101/2024.11.13.24317189

**Authors:** Nadia A. Atai, Kristine Ravina, Saman Sizdahkhani, Robert Rennert, Aidin Abedi, Gavin T. Kress, Federico Fabris, Vincent Nguyen, Stan G Louie, Laura Shin, Isaac Asante, Debra Hawes, Ornella Rossetto, Joseph Carey, Jonathan J. Russin

**Affiliations:** Department of Neurological Surgery and Biomedical Engineering, Neurorestoration Center, Keck School of Medicine, University of Southern California, Los Angeles, CA, USA; Department of Neurosurgery, Carilion Clinic, Virginia Tech Carilion School of Medicine, Roanoke, Virginia, USA; Department of Neurosurgery, Thomas Jefferson University Hospital, Philadelphia, USA; Department of Neurosurgery, University of Utah, Salt Lake City, Utah, USA; Department of Urology, University of Toledo, Toledo, Ohio, USA; Keck School of Medicine, University of Southern California, Los Angeles, California, USA; Icahn School of Medicine at Mount Sinai, New York, NY, USA; Department of Biomedical Sciences, University of Padova, Italy; Department of Neurosurgery, University of Rochester Medical Center, Rochester, NY, USA; Department of Clinical Pharmacy, Mann School of Pharmacy, University of Southern California, Los Angeles, California, USA; Department of General Surgery, Keck School of Medicine, University of Southern California, Los Angeles, California, USA; Department of Pathology and Laboratory Medicine, Children’s Hospital Los Angeles, California, USA; Department of Plastic Surgery, Keck School of Medicine, University of Southern California, Los Angeles, California, USA; Division of Engineering and Applied Sciences, California Institute of Technology, Pasadena, CA, USA

**Keywords:** Artery, Botox, Bypass, Brain, Botulinum toxin A, Spasm, Vasospasm, cSNAP25, ROCK

## Abstract

Vascular graft vasospasm is a lethal risk when using grafts for revascularization and reconstructive surgery. Revascularization is a treatment modality for ischemic diseases including Moyamoya disease that requires bypass surgery. Cerebrovascular graft transplantation carries a 5-10% risk of vasospasm, which can lead to devastating neurological sequelae. Here we report clinical outcomes associated with *ex vivo* botulinum toxin A (BoNT/A) treatment of arterial graft and provide reverse translational studies investigating the potential mechanisms of action of BoNT/A to reduce vasospasm. A retrospective review of the maintained database of patients undergoing surgery was performed for 63 patients. We used paired human vascular graft tissue for *ex vivo* BoNT/A studies to assess for spasmolytic downstream effectors; cleaved SNAP25, pMLC, pMYPT, ROCK1/2, and levels of catecholamines. We found that low-dose BoNT/A graft treatment is associated with 1) a reduction in clinical vasospasm (13.3%, p<0.05) without any identified safety concerns in patients, 2) an increase in arterial cleaved SNAP25, 3) reduced levels of pMLC and pMYPT, and 4) reduced levels of catecholamines. The mechanism of action leading to vascular relaxation is likely through pleiotropic vasodilatory pathways. The application of BoNT/A as a spasmolytic has potentially safe and broad applications across multiple surgical and scientific subspecialties.

## Introduction

Graft vasospasm is a recognized and potentially serious complication after bypass surgeries.^1^ The diagnosis of graft spasm is often made using conventional radiographic imaging modalities in combination with clinical symptoms.^2^ Clinically significant graft spasm was initially described in coronary bypass surgeries.^3, 4^ Revascularization procedures including free flaps, limb or digital replants, extremity bypass, and cerebral bypass have subsequently reported vasospasm as a complication ^5, 6^

Cerebral revascularization is used in the treatment of select complex aneurysms, medically refractory large vessel stenosis/occlusion, Moyamoya disease, and brain tumor resections that require vessel sacrifice.^7, 8^ Autologous arterial interposition grafts are often used for medium- to high-flow extracranial-to-intracranial (EC-IC) or intracranial-to-intracranial (IC-IC) bypass, with arterial grafts generally preferred over venous grafts. This preference is due to higher long-term patency rates and decreased rates of atherosclerosis and intimal hyperplasia, best described in the cardiac literature.^9^ Arterial grafts are at risk for vasospasm, occurring in up to 10% of cases, potentially resulting in disabling ischemia for neurosurgical patients ^10, 11^ To date, the mechanism of graft spasm remains an area of investigation. Based on animal studies it is thought to involve multiple pathways. These include temperature fluctuation/mechanical injury resulting in activation of the ROCK pathway by reactive oxygen species and release of neuromodulators (Norepinephrine, neuropeptide Y) from sympathetic perivascular nerves by intracellular calcium concentration.^10, 12, 13^Unlike the transient calcium fluctuation that initiates vasoconstriction, studies show that the ROCK activation maintains the triggered surge for a longer period of time, suggesting a spatiotemporal interaction between calcium-mediated signaling and the ROCK pathway^14^ Pathological spasm of arterial grafts can lead to graft occlusion and ischemia. While reported strategies for prevention of vasospasm include the use of anti-platelet agents, atraumatic harvest techniques, *ex vivo* treatment with short-acting vasodilators and maintaining an elevated mean arterial pressure (MAP),^15–19^ there is no consensus protocol for spasm prevention.

Botulinum toxin (BoNT) is a neurotoxin produced by several species in the bacterial genus *Clostridium*. It is traditionally classified into seven moieties, four of which (A,B, E, F) can produce toxic reactions in humans.^20–22^ In addition, the A and B varieties are the only type (BoNT/A, BoNT/B) currently used clinically. Indications include the treatment of chronic migraine, spastic disorders, cervical dystonia, hyperhidrosis, frown lines, detrusor hyperactivity^23, 24^ and Raynaud’s phenomenon.^25–28^ Although BoNT primarily works by blocking acetylcholine release at the neuromuscular junction,^20, 29^ pleiotropic effects have been suggested in pre-clinical studies.^29–32^

Our team previously demonstrated *ex vivo* BoNT application for off-label use to prevent graft spasm in revascularization,^29, 33, 34^ theoretically reducing the risk of systemic effects of BoNT such as anaphylaxis, respiratory insufficiency, and generalized muscle weakness.^35, 36^ The method previously described for prevention of vasospasm with BoNT/A is to incubate the graft in reconstituted BoNT/A^29, 33^. However, this use is off-label and to our knowledge, no prior reports have described BoNT/A efficacy and safety related to human cerebral revascularization. In addition, a knowledge gap exists in understanding the pleiotropic effect of BoNT/A that may contribute to spasmolysis in human arteries.

We herein report the use of BoNT/A in 63 patients undergoing cerebral revascularization surgery and describe the clinical outcomes compared to historical controls. In addition, we use a reverse translation approach in human tissue specimens to investigate the mechanism of action of BoNT/A’s effect on arterial graft reactivity, focusing on adrenergic signaling and the Rho/ROCK pathway for smooth muscle contraction.

## Results

### Characteristics of study participants

Sixty-three patients (35 women [55%]; mean age 50.76±14.06 years, Table 1) underwent cerebral revascularization surgery, with a total of 66 grafts in 63 patients. BoNT/A was used in 35 patients (37 grafts [56%]). Median follow-up time was 175 days (interquartile range: 36-936). Indications for bypass included ruptured aneurysm (35%), ischemia (23%), unruptured aneurysm (21%), moyamoya disease (16%), and tumor (5%) (Fig. 1 and Table 1). Twenty-five patients presented with strokes, 17 of which were acute. While most characteristics had no significant differences, notably, the time of the last follow-up differed significantly at p < 0.001. In addition, BoNT/A patients were significantly (p = 0.007) more likely to have presented with a recurrent stroke, however, there was no significant difference in the prevalence of recurrent stroke if a stroke was present. BoNT/A patients were significantly less likely to present with an aneurysm (p = 0.004) but more likely to present with ischemia (p = 0.011).

**Table 1.**
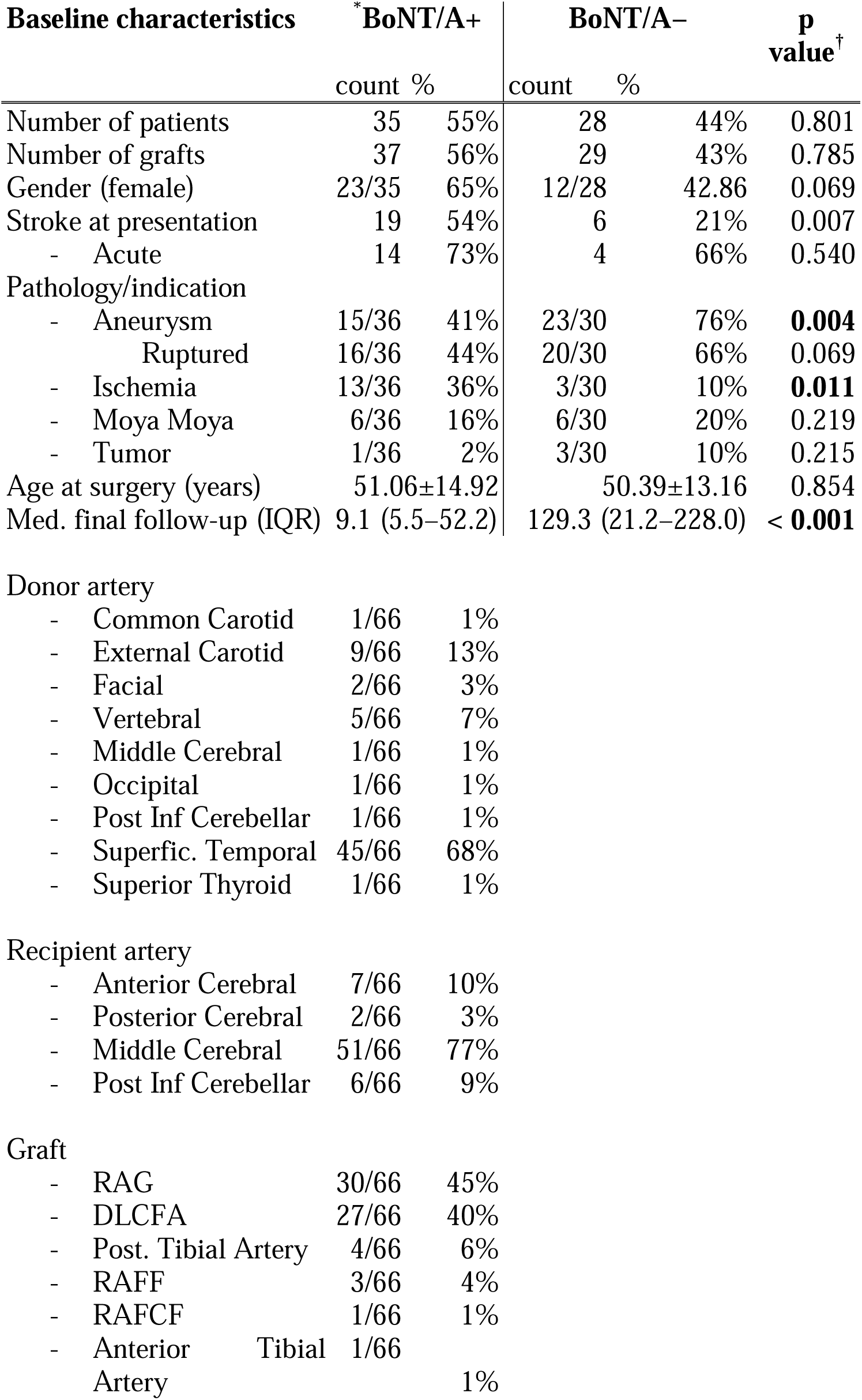
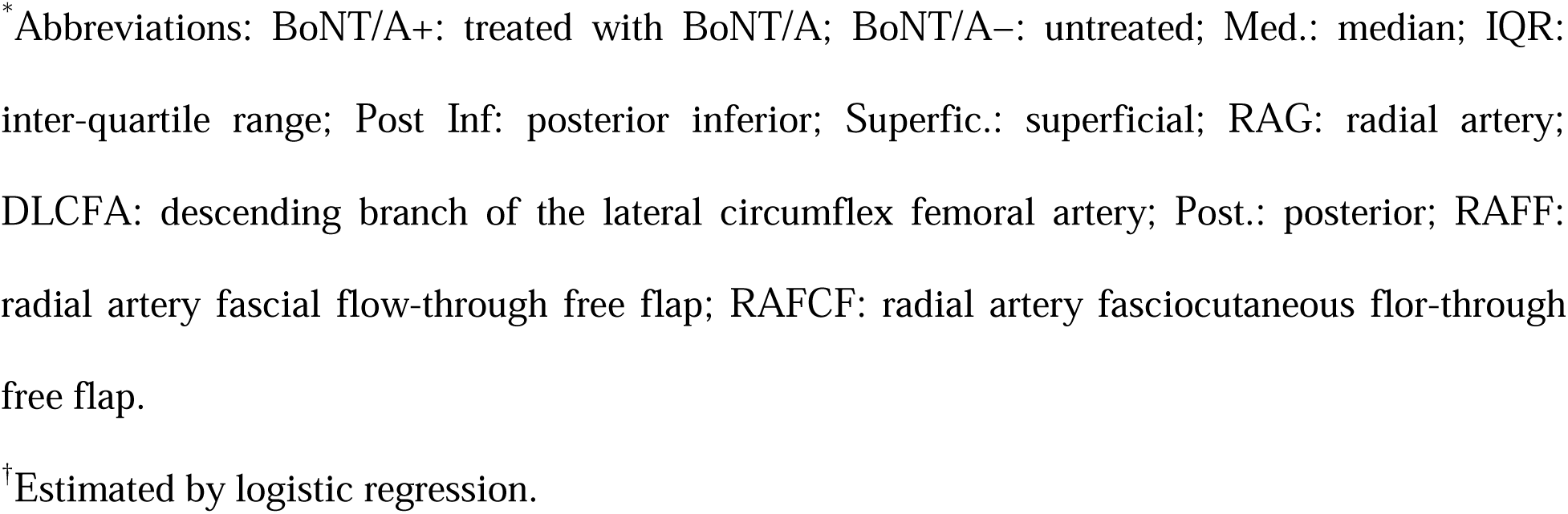
Baseline characteristics of study participants.

**Fig. 1.**
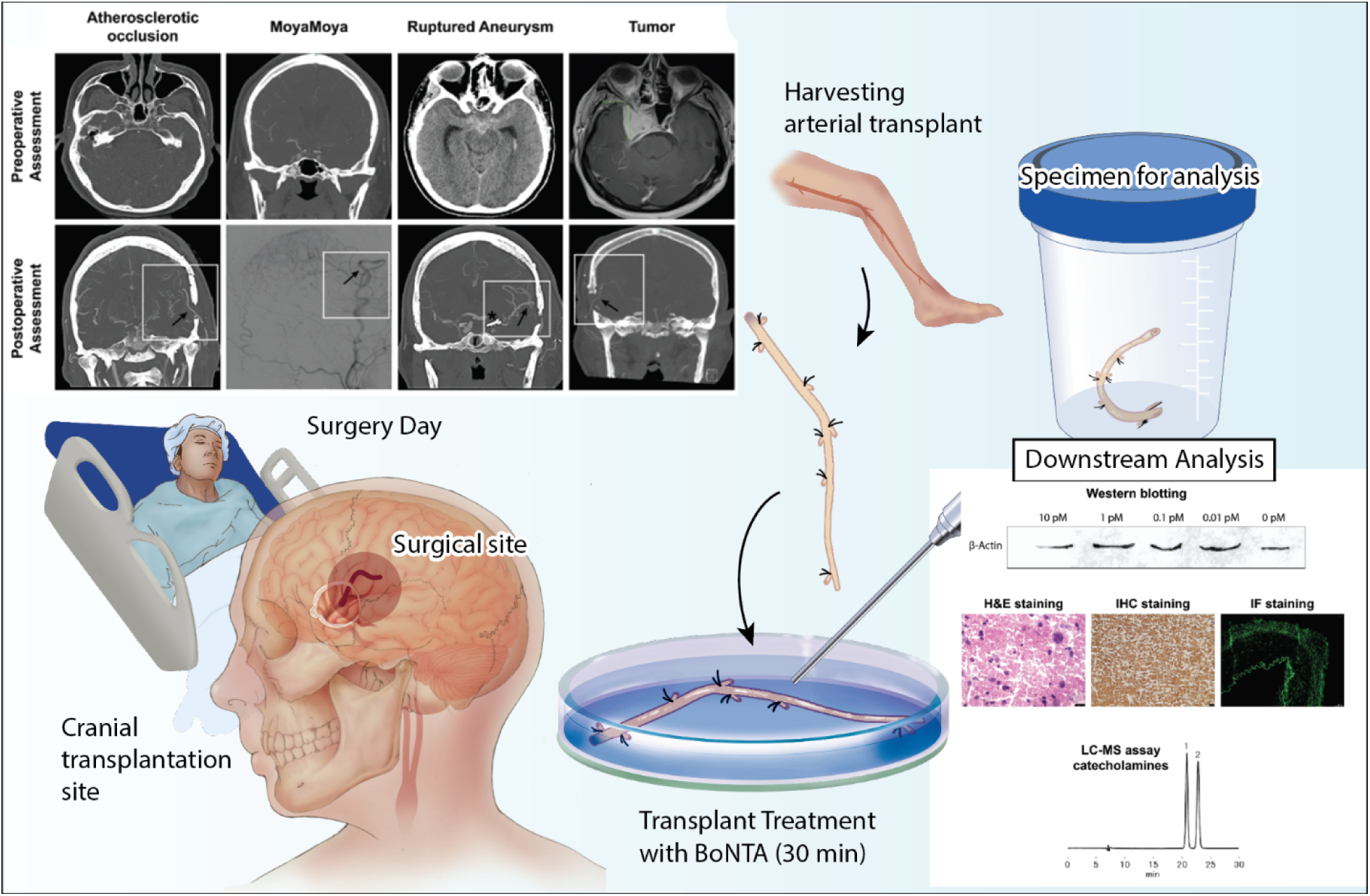
Illustration of identification of stroke population with various etiology using clinical diagnostic characteristics and current workflow for downstream experimental assays. Clinical image of selected patients with various surgical indications. The superior images show preoperative assessment of the brain pathology and inferior images depict BoNT/A treated graft patency after surgical procedure. The arrow indicates the arterial graft used in the corresponding patient. Star indicates an intracranial permanent clip used for an intracerebral aneurysm. Workflow of the surgical procedure; Upon admission, the patient undergoes a multidisciplinary diagnostic assessment, and surgery is planned based on the pathology. 1) The neurosurgeon identifies the location (surgical site) of the graft transplantation in the brain, 2) the plastic microsurgeon identifies the potential arterial segment to harvest and treats with BoNT/A. After 30 minutes in BoNT/A (control saline), the neurosurgeon measures the length required for the surgical transplantation and the remaining unused tissue is collected for downstream analysis in the lab. In the lab, the arterial specimen is aliquoted and processed for various downstream processes to assess the mechanism of action of BoNT/A in arterial graft patency. This assay includes at the protein level; Western Blotting (WB), H&E, immunohistochemistry staining (IHC), immunofluorescence staining (IF), at molecular levels; Liquid chromatography-mass spectrometry (LC-MS) assay.

### Vasospasm, stroke, mortality, and functional outcomes

The incidence of clinically significant graft vasospasm (Fig. S1) was significantly lower in the BoNT/A versus control groups (0 vs. 13.3%, *p*=0.040). Rates of angiographic spasm were not significantly different between the BoNT/A and control groups. Inter-observer reliability was low (kappa 0.52 for dichotomous assessment and 0.36 for the four-point scale), and when the ratings from each observer were analyzed individually, there was no significant difference between the two groups. In addition, multinomial logistic regression of discharge disposition versus BoNT/A treatment revealed that BoNT/A treatment was associated with significantly (p = 0.014) lower discharge rate directly home (17.82% vs. 56.61%). The two groups were comparable in terms of 30-day all-cause stroke rate (*p*=0.951), 30-day all-cause mortality (*p*=0.473), post-operative graft patency (*p*=1.0), length of hospital stay (*p*=0.756), and improvement in mRS from baseline to final follow-up (*p*=0.482). A model that considered the different follow-up times for BoNT/A and control patients did not find a significant difference for mRS.

### BoNT/A induced cleavage of SNAP25 in human brain samples

An antibody specific to the BoNT/A cleavage product of SNAP25 (Fig. 2A) was used to probe western blotting of human brain tissues (Fig. 2. B, Fig. S4-S5). This produced a band at the expected kDa as well as a doublet in the cleaved human product. Immunohistology with the cSNAP25 antibody (Fig. 2C) revealed a visible increase in cleaved SNAP25 after BoNT/A treatment, as expected. Densitometry of western blotting (Fig. 2D) showed that BoNT/A treatment resulted in increased measured cleaved SNAP25 (cSNAP25), which was significant, with a non-significant decrease in full-length SNAP25. The doublet band in human tissue cleavage product has been reported and characterized elsewhere, representing cleaved (24kDa) and non-cleaved SNAP25 (25kDa) depending on the amount of neurotoxin used.^41^

**Fig. 2.**
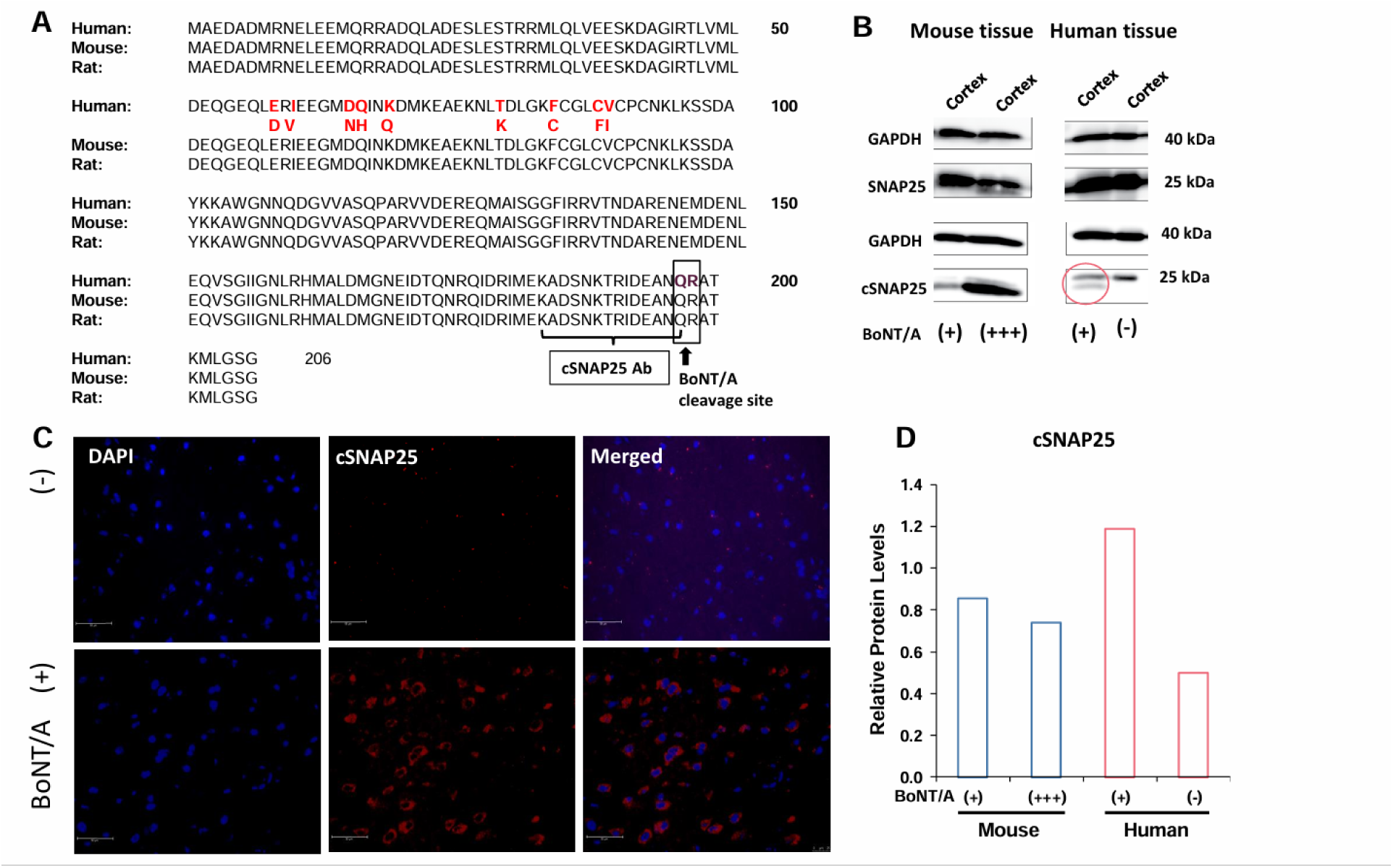
Validation of antibody to mouse cleaved SNAP25 for measurement of human SNAP25 cleavage by BoNT/A. A) Alignment of Mouse and Human SNAP-25 protein sequenced, indicating site of BoNT/A cleavage of the target protein. Red letters show specific amino acid differences between human SNAP25a and SNAP25b splicing variants. B) Western blot of BoNT/A treated (+) and untreated (-) human tissue and mouse tissue with low (+) and high dose (+++) BoNT/A. A double band in the BoNT/A treated tissue is indicated. C) Immunofluorescence of human brain tissue before and after treatment with BoNT/A, probed for cleaved SNAP25 and DAPI. Note the concentration of cleaved SNAP25 on cell peripheries. D) Densitometry of GAPDH-adjusted western signal. The graph is for one point per treatment and is a qualitative presentation of Fig. 2B.

### BoNT/A cleavage of human arterial SNAP25

Upon treatment of arterial graft material with BoNT/A, we extracted total protein and performed western blotting (Fig. 3A). We did not quantify these blots because of known wide variations of nerve distribution in arterial tissue segments and attendant difficulties in normalizing with conventional housekeeping genes such as GAPDH or β-actin within biologically different individuals. While each sample could be normalized to its own housekeeping genes, these would not have been comparable to other biological replicates because of both nerve distribution and heterogeneity of cell activity. However, in those arterial samples that had uniform morphology (Fig. S6) and SNAP25, we subjected both treated and untreated samples and corresponding controls (Fig. 3B-C) to immunoassaying for cleaved SNAP25, TH, and DAPI (Fig. 3 B-C). Assessing the percent of cells with cleaved SNAP25 signal (Fig. 3D), revealed a mean 14.52% ± 5.52% SEM presence of cells showing cleaved SNAP25 colocalizing with TH. This finding was significant (p < 0.001, *R*^2^ = 0.600).

**Fig. 3.**
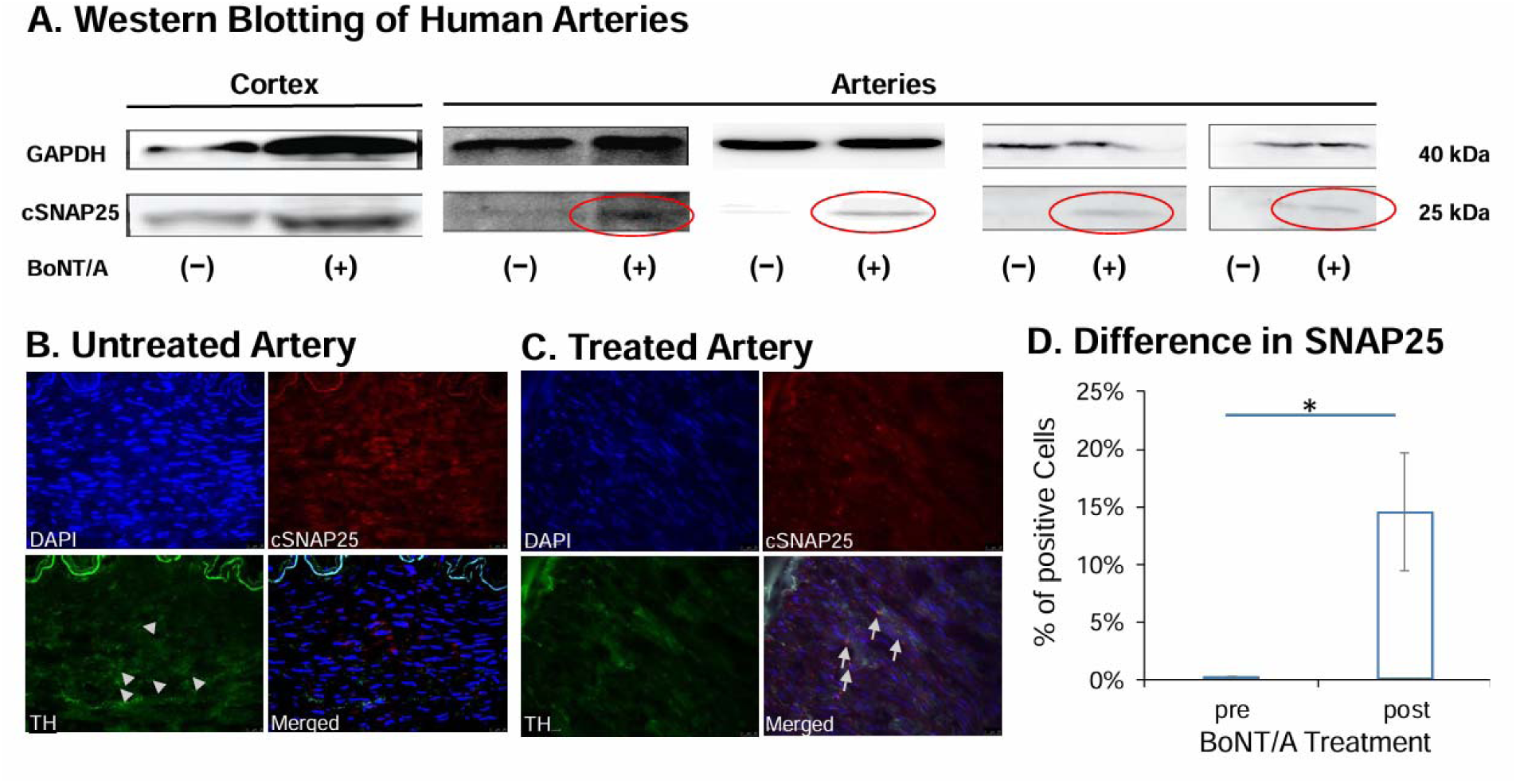
Western blotting and quantitative immunofluorescence of effects of BoNT/A treatment in human arteries. A) Qualitative western blotting of human cleaved SNAP25 in untreated and BoNT/A treated brain cortex and arterial samples. B) Immunofluorescence of representative human artery without BoNT/A treatment. C) Immunofluorescence of the human artery with BoNT/A treatment. D) Analysis of cleaved SNAP25 in pre-vs. post-treated arteries revealed a significant increase in cleaved SNAP25 after treatment (n=5, p< 0.05).

### BoNT/A reduces levels of major neurotransmitters in human arteries

When we exposed tissue samples to BoNT/A and measured neurotransmitter levels in arteries with and without exposure by LC-MS, we found that BoNT/A treatment was associated with a reduction in multiple (but not all) neurotransmitters in the panel (Fig. 4A-G). Specifically, when analyzed by paired generalized linear mixed models, with random intercepts for each donor, we found that BoNT/A administration significantly (p ≤ 0.05) reduced levels of norepinephrine, epinephrine, dopamine, glutamine, and 5HIAA. Reduction was between 33% to 65% of untreated levels (Table 2). Glutamate and VMA level alterations were not significant.

**Fig. 4.**
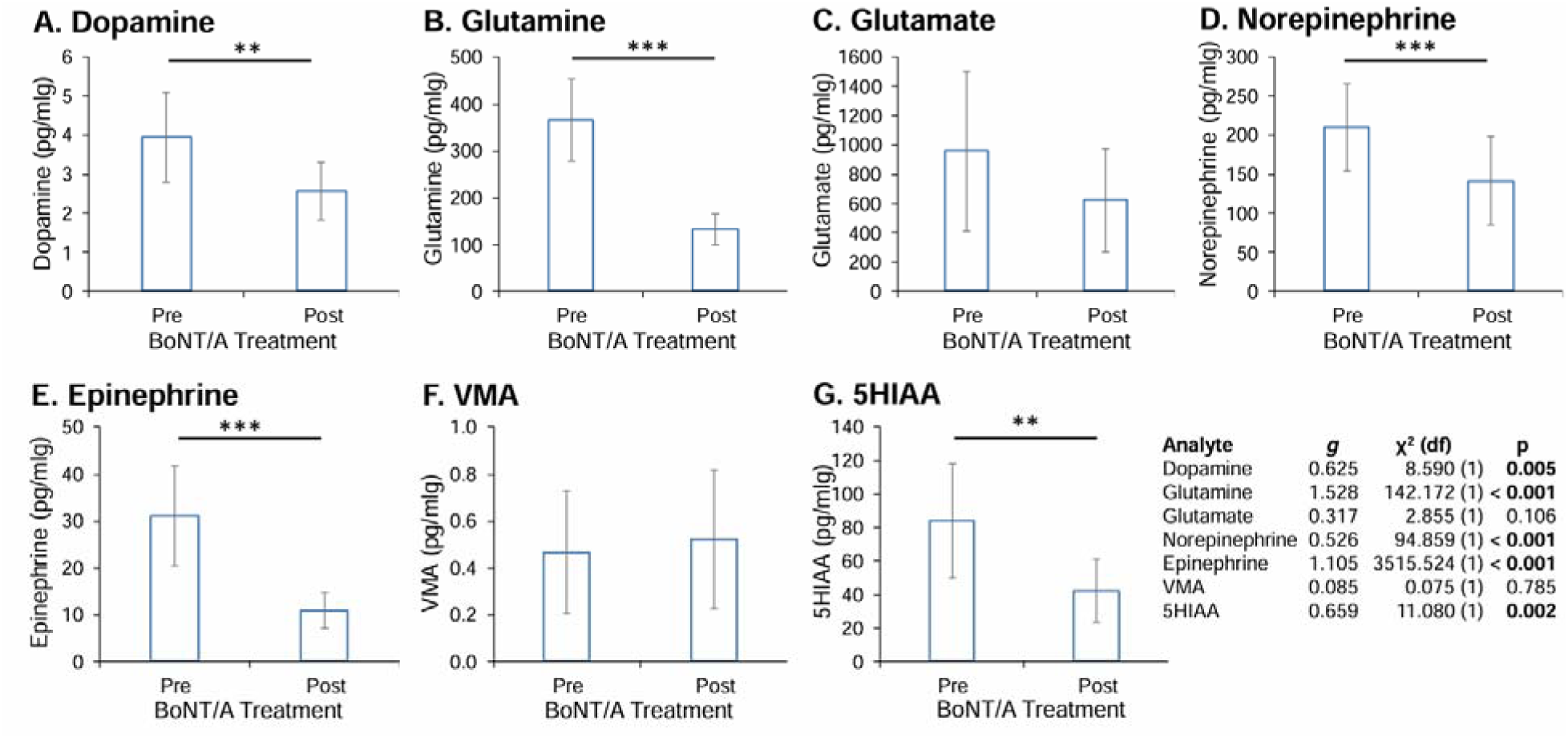
LC-MS of selected neurotransmitters before and after treatment by BoNT/A. Arteries were harvested, and adventitia were removed and subsampled for LC-MS. Next. arteries were treated with BoNT/A (n=4), and treated arteries subjected to LC-MS, all as described herein. A) Glutamine. B) Glutamate. C) Norepinephrine. D) Epinephrine. E) VMA. F) 5HIAA. Analysis revealed significant changes in several of the neurotransmitters.

**Table 2.** Arterial neurotransmitter level changes induced by BoNT/A.

| Analyte | BoNT/A+<sup>*</sup> |  | BoNT/A– |  | % Change | g<sup>†</sup> | χ<sup>2</sup> (df) | p<sup>‡</sup> |
| --- | --- | --- | --- | --- | --- | --- | --- | --- |
| Dopamine | §3.955 | ±1.140 | 2.572 | ±0.742 | –35% | 0.625 | 8.590 (1) | **0.005** |
| Glutamine | 366.774 | ±88.092 | 132.462 | ±33.395 | –64% | 1.528 | 142.172 (1) | < **0.001** |
| Glutamate | 957.962 | ±543.589 | 623.482 | ±354.553 | –35% | 0.317 | 2.855 (1) | 0.106 |
| Norepinephrine | 210.011 | ±56.516 | 141.592 | ±56.516 | –33% | 0.526 | 94.859 (1) | < **0.001** |
| Epinephrine | 31.078 | ±10.601 | 10.867 | ±3.711 | –65% | 1.105 | 3515.524 (1) | < **0.001** |
| VMA | 0.466 | ±0.261 | 0.520 | ±0.297 | +12% | –0.085 | 0.075 (1) | 0.785 |
| 5HIAA | 84.119 | ±34.426 | 42.022 | ±18.835 | –50% | 0.659 | 11.080 (1) | **0.002** |

<sup>\*</sup>Abbreviations: BoNT/A+: treated with BoNT/A+; BoNT/A–: untreated; VMA: vanillylmandelic acid; 5HIAA: 5-hydroxyindoleacetic acid.
<sup>†</sup>Hedge's *g* standardized difference.
<sup>‡</sup>Adjusted by Benjamini-Hochberg FDR for 7 simultaneous tests.
<sup>§</sup>pg/mg tissue, estimated from models.

### BoNT/A alters ROCK pathway proteins in treated arteries

We performed western blotting on Rho kinase (ROCK) and its downstream targets: phosphorylated myosin phosphatase target subunit 1 (MYPT1), and phospho-myosin light chain 2 (pMLC) in arterial graft samples collected before and after BoNT/A treatment (Fig. 5A-F, Table 3). We determined that treatment was associated with a reduction in all three proteins measured but found that the reduction of ROCK was not significant. We note that the data was generated with two different housekeeping proteins for normalization to address the possible effects in a supplement.

**Fig. 5.**
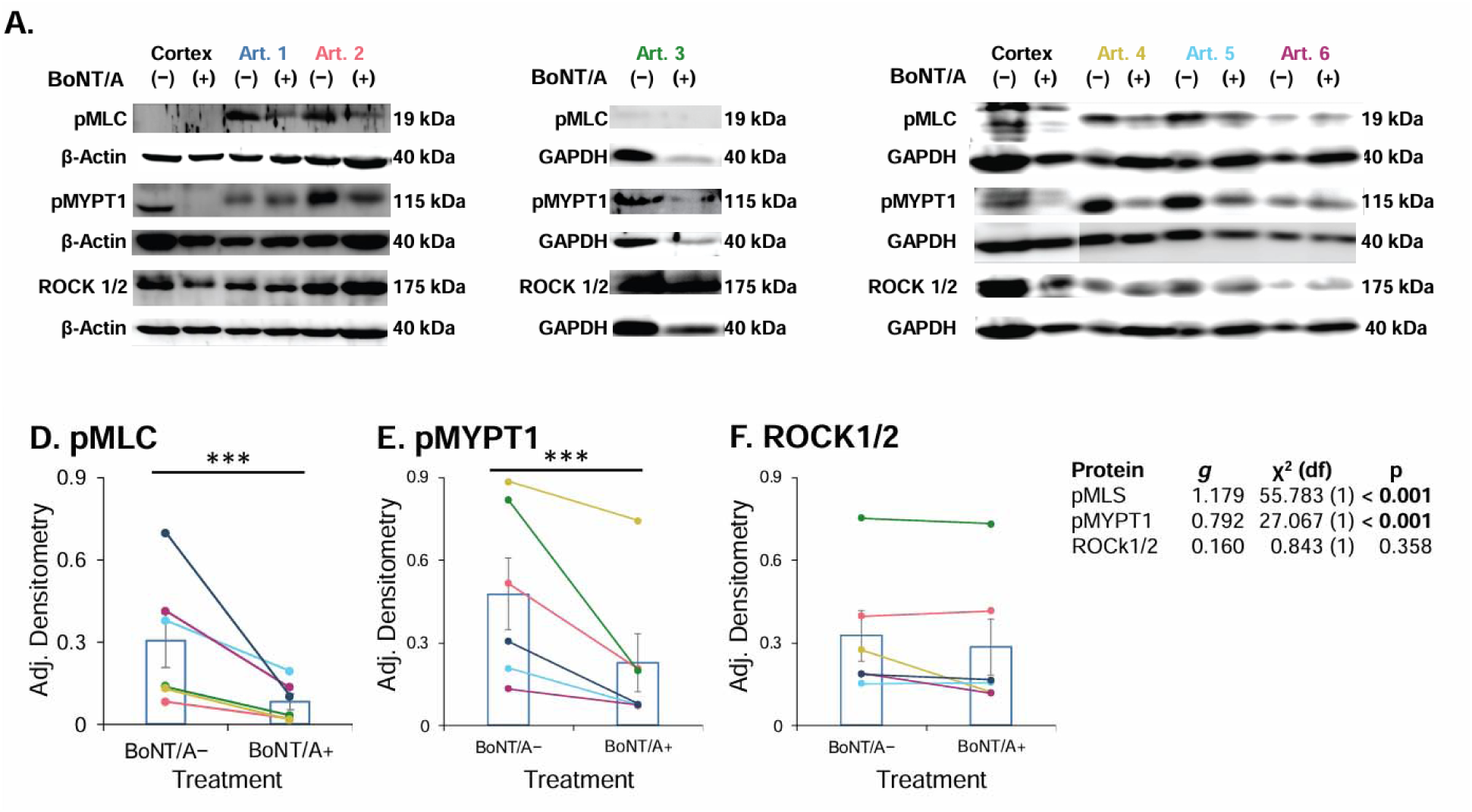
Western blot of pMLS, pMYT1, and ROCK1/2. Western blot of arterial samples with and without BoNT/A treatment, probed with antibodies for A) pMLS, B) pMYT1, and C) ROCK1/2, as well as GAPDH. Analysis of GAPDH adjusted densitometry for D) pMLS, E) pMYT1, and F) ROCK1/2 revealed a trend of reduction with BoNT/A treatment, although the ROCK1/2 trend was not detectable significant (n=6).

**Table 3.** Arterial ROCK pathway protein level changes induced by BoNT/A.

| Protein | BoNT/A+<sup>*</sup> |  | BoNT/A– |  | % Change | g<sup>†</sup> | χ<sup>2</sup> (df) | p<sup>‡</sup> |
| --- | --- | --- | --- | --- | --- | --- | --- | --- |
| pMLC | §0.305 | ±0.096 | 0.082 | ±0.029 | –74% | 1.179 | 55.783 (1) | < **0.001** |
| pMYPT1 | 0.477 | ±0.129 | 0.228 | ±0.106 | –56% | 0.792 | 27.067 (1) | < **0.001** |
| ROCK1/2 | 0.325 | ±0.093 | 0.284 | ±0.100 | –17% | 0.160 | 0.843 (1) | 0.358 |

<sup>\*</sup>Abbreviations: BoNT/A+: treated with BoNT/A+; BoNT/A–: untreated; pMLC: phosphorylated myosin light chain; pMLC: phosphorylated myosin phosphatase; ROCK1/2: rho-associated protein kinase.
<sup>†</sup>Hedge's *g* standardized difference.
<sup>‡</sup>Adjusted by Benjamini-Hochberg FDR for 3 simultaneous tests.
<sup>§</sup>Densitometry, estimated from models.

## Discussion

Cerebral revascularization using arterial interposition grafts is an important strategy for the treatment of complex ischemic and non-ischemic vessel pathologies.^1–5^ Limb arteries such as the radial artery are frequently used as interposition grafts due to ease of access and harvest. These arteries, however, are classified as high-risk “spastic” due to a predominance of vasoconstrictor receptors, relatively reduced production of vasodilators, and an abundance of vascular smooth muscle cells (VSMCs).^6,7^ Early cerebral revascularization interposition graft occlusion and spasm occurs in approximately 5-10% of cases despite advancements in surgical technique and pharmacologic treatment.^4,8,9^ Therefore, revascularization graft vasospasm resulting in recurrent ischemia and/or occlusion remains a potentially lethal complication.^7,10,3,4,8^

Various factors that alter vasomotor tone can provoke arterial narrowing, which can restrict or prevent blood flow. In physiological conditions, arteries can modify their diameters through corrective dilation or constriction of the artery to condition the blood supply to vital tissues. This autoregulation system is challenged in pathologic conditions, such as Prinzmetal’s Angina, Raynaud’s Disease, Systemic Sclerosis, Chronic Kidney Disease, Multiple Sclerosis, Buerger’s Disease, preeclampsia, and chemo/radiation-induced vasospasm.^42, 43^ Moving beyond disease context, certain surgeries are intricately linked with a similar mechanism of action of arterial spasm including but not limited to, cerebral bypass, coronary bypass, carotid bypass, renal bypass, hepatic bypass, traumatic spinal cord bypass, infrainguinal bypass, and free flap transplantation in reconstruction surgeries (Table 4).^15, 44–46^ Arterial spasm in the context of bypass surgery is a feared complication that arises from a complex interplay of factors between the recipient and donor artery in combination with the vessel microenvironment due to the manipulation and clamping of blood vessels during surgery. This can induce postoperative inflammation and damage to the endothelial lining of blood vessels, and temporary cessation of blood flow and subsequent reperfusion further heightens the spastic risk.^47^ The rate of this complication in all bypass surgeries is low, but the mortality can be high.^48^

**Table 4.** Surgical Vasospasm indications that may benefit from BoNT/A graft pre-treatment.

| **Indication** | **Type(s) of Vessel** | **Primary Patency Rate** | **Common Causes of Reduction of Patency** |
| --- | --- | --- | --- |
| Cerebral Revascularization<sup>65</sup> | Radial Artery | 94% | Stroke/Vasospasm |
|  | Saphenous Vein | 93% | Stroke |
| Coronary Artery Bypass<sup>66, 67</sup> | Radial Artery | 98% | Thrombosis, Vasospasm |
|  | Saphenous Vein | 86% | Thrombosis |
| Raynaud's Syndrome<sup>68</sup> | Varies | 83% | Thrombosis |
| Free Flap<sup>6</sup> | Varies | 82% | Infection, Thrombosis, Technical Error |
| Digital Replant<sup>69</sup> | Varies | 63% | Technical challenges associated with the site/mechanism of injury |
| Infrainguinal Bypass<sup>70</sup> | Femoral artery | 63% | Neointimal Hyperplasia, Thrombosis, Technical Error |
| Revascularization for Traumatic Injury<sup>71,72</sup> | Varies | 93% | Technical error, Thrombosis |

Primary pharmacologic treatment for vasospasm is currently calcium channel blockers (CCBs), notably verapamil or diltiazem, serving as frontline agents to induce smooth muscle relaxation in cerebral and coronary arteries. Sublingual nitroglycerin, a vasodilator, plays a pivotal role in acute coronary spasm management, with long-acting formulations prescribed for sustained relief. Isosorbide dinitrate or mononitrate represents an additional avenue for vasodilation, while beta-blockers, cautiously administered, may be considered to alleviate arterial workload.^47^ Other treatment strategies for vasospasm include intra-arterial drug delivery and mechanical angioplasty as salvage treatments for graft vasospasm.^8^ In cases where these therapies fall short, a repeat surgical intervention for graft revision can be attempted but this subjects the patient to accumulating risks and can prolong hospitalization.^8,11^ Moreover, the need for blood pressure maintenance in the postoperative period for cerebrovascular bypass patients generally limits systemic vasodilator therapy.^5^ Additionally, intraoperative preventive and post-operative salvage vasodilator effects are typically short-lived.^12^ Although different variations of spasmolytic interventions have been proposed for vasospasm in various diseases and surgical procedures, there currently exists no single effective spasmolytic agent or protocol for graft vasospasm prevention.^7,10^ Recognizing these treatment-induced limitations are pivotal for anticipating and managing vasospasm in the context of various bypass surgeries while integrating pharmacologic treatments.

In this study, we assessed the effects of *ex vivo* graft treatment with BoNT/A for preventing graft vasospasm following cerebral revascularization surgery. As previously described, BoNT primarily works by blocking acetylcholine release at the neuromuscular junction and is used for multiple medical conditions including headache prophylaxis, urinary incontinence, hyperhidrosis, hemifacial spasm, spasticity disorders, and dystonia. ^20, 29^ While hazards are associated with BoNT/A use, these are primarily due to high dose, frequent use, and prolonged exposure causing retrograde axonal migration of BoNT from the site of injection.^35, 36^ BoNT/A can prevent arterial spasms in animal pre-clinical studies.^29–32^ A recent systematic review and meta-analysis of controlled animal studies in 1032 animals with various doses of BoNT/A showed that BoNT/A increases vasodilation in arteries by 40% and in veins by 43% while decreasing the risk of thrombosis (arteries 85% and veins 76%).^49^ The authors concluded that intraoperative application or pre-treatment with BoNT/A alters the vascular system, suggesting an expansion of research and clinical use of BoNT/A in human vascular disease and surgery.

Our group reported in 2018 the first 3 human cerebrovascular cases receiving intraoperative BoNT/A for prevention of cerebrovascular graft vasospasm, demonstrating feasibility.^34^ In the current study we report a significant decrease in the incidence of clinical vasospasm in the BoNT/A+ group vs controls. No difference was identified in 30-day stroke, all-cause mortality, or long-term functional recovery as measured by mRS. These findings have significant implications for the safe performance of revascularization procedures. Our clinical data analysis did not reveal a significant difference in angiographic vasospasm rates with BoNT/A graft preparation. This was likely influenced by the poor inter-observer reliability we encountered, an issue previously described in the assessment of vasospasm following SAH.^50^ This decoupling of angiographic and clinical outcomes is also consistent with prior data on the administration of the calcium channel blocker nimodipine for clinical vasospasm prevention (one of few therapies shown to improve outcomes after aneurysmal subarachnoid hemorrhage).^51–53^ Although acting via a different mechanism than that posited for BoNT/A (calcium channel blockade reduces post-synaptic cell contractility), the effect of nimodipine on rates of radiographic vasospasm is inconsistent, with pleiotropic effects hypothesized outside of direct effects on large-vessel reactivity.^54–56^ Reconciling the notion that BoNT/A primarily acts via pre-synaptic release of acetylcholine while graft spasm is likely largely adrenergic, i.e., norepinephrine release,^29, 33^ our results support that BoNT/A acts through pleiotropic vasodilatory pathways to mitigate clinical vasospasm.

Our clinical study was “reverse translated” to evaluate the pleiotropic effects of BoNT/A on human arterial tissue. A total of 29 human arterial graft samples matched per patient (non-BoNT/A and BoNT/A arterial segments), were included in this study. We found that BoNT/A treatment resulted in cleavage of SNAP25 in the graft tissue. Despite quantification being difficult due to the low concentration of BoNT/A used, low levels of SNAP25 in some arteries, and heterogenous distribution of nerve terminals, co-localization and quantification of cleaved SNAP25 was possible in peri-arterial pre-synaptic nerve terminals. Fitting with a decreased pre-synaptic release of these transmitters from SNAP25 inhibition, BoNT/A treatment was associated with reduced levels of several neurotransmitters from sympathetic perivascular nerves in arterial tissue, except glutamate and VMA. The levels of phosphorylated myosin light chain and phosphorylated myosin phosphatase were also significantly reduced. Although the kinase ROCK1/2 was overall reduced, this change was not detectable significant.

These findings suggest BoNT/A has an impact on several convergent pathways related to vascular constriction (Fig. 6). The catecholamines, dopamine, norepinephrine, and epinephrine play important roles in sympathetic nervous activity to induce physiologic vascular constriction, however, these signaling pathways can become hyperactive under pathologic conditions and cause cerebral infarctions (i.e., after aneurysmal subarachnoid hemorrhage).^57, 58^ Currently, vascular graft spasm is thought to result in parts from catecholamine release by denervated nerve endings in the harvested tissue into the perivascular space, which then activates a PLC-β/IP_3_/Ca^2+^ contraction pathway via MLC phosphorylation. ^59^The demonstrated cleavage of SNAP25 by BoNT/A resulting decrease in sympathetic perivascular catecholamine levels provides one mechanistic explanation for the reduction in clinical vasospasm seen in this human trial (Fig. 6). Modulation of the separate but convergent vasoconstrictive ROCK signaling pathway, activated by cold and mechanical stimuli, is also suggested by our data. This pathway is likely contributory to vasospasm as the OR room temperature is maintained at 25°C, as compared to physiologic body temperature of ∼37°. This relative cold can induce ROS, which can activate ROCK. Additionally, mechanical stimulus, such as graft harvest, can induce activation of the upstream signaling molecule RhoA, also contributing to ROCK activation.^14^ The decrease in pMYPT1, a key molecule in the ROCK pathway, shown in this work provides an additional mechanistic explanation for our clinical findings. Finally, the catecholamine and ROCK pathways merge at the effector molecule MLC and inhibition of MLC phosphorylation has been shown to prevent vasospasm.^60^ The decrease in pMLC seen with BoNT/A application in this work is consistent with a convergent treatment effect on these pathways. Additional exploration of alterations in these and other pathways may nonetheless lead to a more refined understanding of the mechanisms of action. Future applications of BoNT/A to reduce post-operative graft vasospasm and broader applications will likely not be limited to cerebrovascular.

**Fig. 6.**
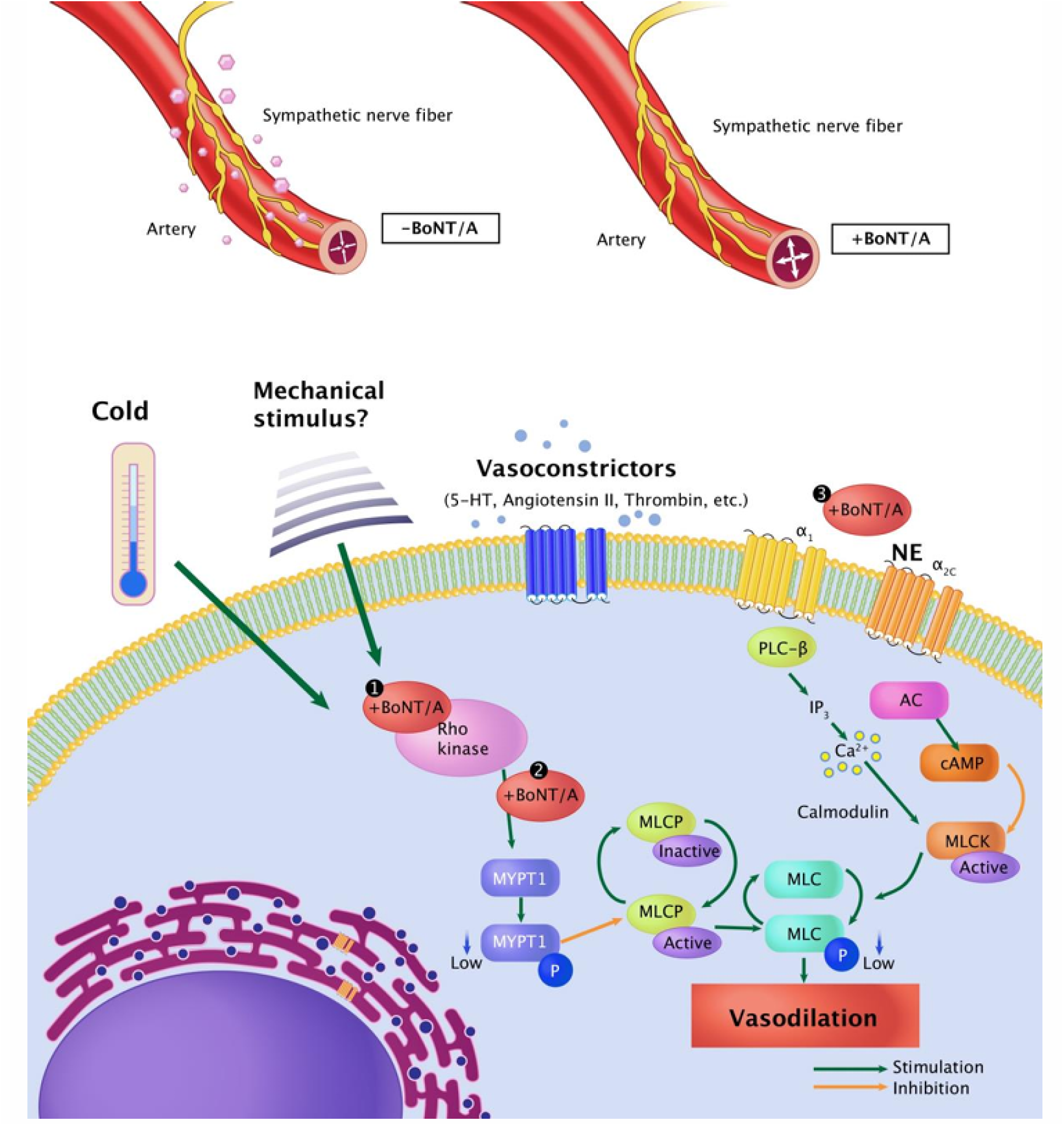
Models of spasmolytic pathways induced by BoNT/A to reduce graft vasoconstriction. BoNT/A inhibition of adrenergic and ROCK pathways in vascular smooth muscle cell (VSMC) activity. Adrenergic VSMC contraction occurs via two pathways depending on which receptor is activated. Catecholamines binding to the α_1_ receptor activates phospholipase C-β (PLCβ), which increases intracellular Ca^2+^ levels mediating MLCK activation. NE binding to the α_2_ receptor inhibits adenylyl cyclase (AC), subsequently preventing inactivation of MLCK. This results in phosphorylation of the MLC by MLC kinase (MLCK) to mediate VSMC contraction. The ROCK pathway is primarily activated by vasoconstrictive substance binding to a G protein-coupled receptor and activating guanine nucleotide exchange factor (GEF). It can also be activated by reactive oxygen species (ROS) produced by the VSMC mitochondria upon cold exposure or via mechanical stimulus. VSMC contraction can also be promoted by MLCP inactivation by MYPT1, a ROCK downstream effector. Our current study shows that BoNT/A-mediated (STX A) vasodilation occurs via the inhibition of multiple synergic pathways. This includes 1 and 2) ROCK pathway inactivation via reduced phosphorylation of downstream effector MYPT1 (MYPT1p)/MLCP and 3) inhibition of catecholamines release (Nor/Epi) via cSNAP25, in perivascular space to prevent adrenergic receptor activation and corresponding downstream pathways in VSMCs.

There are significant limitations to this study. To our knowledge, this is the first in human study investigating the transplantation of arterial grafts exposed to BoNT/A intracranially. Given the novelty of the approach, we were without any prior standardized tissue processing protocol. In addition, this was a single-center, non-randomized retrospective design with limited access to human tissue. Human specimens were limited by clinical availability and at times the tissue volumes were not ideal for mechanistic evaluations and parallel studies with various methods. Safety concerns also limited our protocol to a single low dose (10 Units/mL) and operative workflow resulted in short exposure (30 minutes) of BoNT/A.

## Conclusions

The treatment of human arterial grafts with BoNT/A prior to intracranial transplantation has a favorable safety profile and reduced rates of clinical vasospasm. A reverse-translational model demonstrates BoNT/A has a pleiotropic mechanism that cleaves pre-synaptic SNAP-25, reduces catecholamine levels, and modulates the ROCK pathway.

## Methods

### Retrospective Study Population

An IRB-approved, prospectively maintained database was used to identify and consent patients who had undergone EC-IC bypass with an autologous arterial interposition graft between July 2014 and January 2022. Interposition grafts were treated with BoNT/A beginning in March 2017. Information obtained included patient demographics, clinical presentation, indication for bypass, surgery performed, pre- and post-operative imaging characteristics, clinical outcomes (e.g., modified Rankin Scale [mRS] at follow-up), and complications.

### Surgical Protocol

All procedures were performed at a single institution by a multidisciplinary surgical team including neurosurgery and plastic/reconstructive surgery. Patients were considered candidates for cerebral revascularization after failing conservative treatment or if lesions were not amenable to traditional microsurgical or endovascular approaches based on multidisciplinary review. The revascularization strategy (Fig. 1) was planned through collaboration with the plastic and neurosurgical teams. The donor vessel was harvested by the plastic surgery team in all cases. Donor vessels included the radial artery (RAG), radial artery fascial flow-through free flap (RAFF), radial artery fasciocutaneous flow-through free flap (RAFCF), descending branch of the lateral circumflex femoral artery (DLCFA), and anterior and posterior tibial arteries. Each graft was treated *ex vivo* with BoNT/A (Botox, Onabotulinumtoxin A, Allergan Incorporated, Irvine, California), as described herein, before implantation.^63^ All grafts were harvested in a minimal traumatic manner. The adventitia was removed, and the graft was irrigated and incubated in 100 units of BoNT/A in 10 mL of sterile normal saline for approximately 30 minutes. Prior to transplantation, it was irrigated with a heparin/milrinone saline solution (10,000 units of heparin with 10 mg of milrinone in 1 liter of normal saline).

### Outcome assessment

Outcomes assessed included post-operative radiographic and clinical vasospasm, 30-day all-cause stroke rate, 30-day all-cause mortality, post-operative graft patency, length of hospital stay, hospital discharge, and changes in modified Rankin Scale (mRS) from baseline to final follow-up. Radiographic graft vasospasm (Fig. S1) was rated on a dichotomous (present/absent) and a four-point scale (no vasospasm, mild, moderate, or severe vasospasm) based on a blinded review of angiographic images by four observers (including two dual-trained vascular neurosurgeons, one open vascular neurosurgeon, and one neuroradiologist; excluding the treating surgeon). Clinical graft vasospasm was defined as any new neurologic deficit that correlated with angiographic spasm of the bypass graft’s territory and was not otherwise explained. This was assessed independently from vasospasm secondary to subarachnoid hemorrhage (SAH).

### Assessing BoNT/A substrate, synaptosomal-associated protein 25 (SNAP-25) cleavage in perivascular adrenergic nerve terminals

Sequential 4-µm-thick surgical sample sections were prepared from human brain and artery FFPE tissues for immunohistochemistry (IHC, Fig. S2) and immunofluorescence (IF) staining. Samples were carefully selected based on the matching size of tissue and intact morphology. Chromogen-based IHC analysis for BoNT/A and full-length SNAP25 was performed by using an automated staining system (BOND-MAX; Leica Microsystems, Vista, CA). For IHC analysis, antibodies were titrated on the basis of the minimum to maximum range of staining negative to positive in the control human brain specimens. Briefly, slides were fixed with a 16% PFA (EM grade): PBS/dH_2_O at a 1:2:1 ratio for 10 minutes, followed by 3x TBS/dH_2_O resin and soaked in Bond buffer. Slides were loaded in Bond Rx autostainer, ER-1-20m (pH6 retrieval). Rabbit primary Ab for full-length SNAP25 was applied at 1:200 in artery samples and 1:5000 in brain tissue for 45m. Secondary Ab E41-20m, Rb DAB for 30m. Slides were unloaded, and coverslips were applied with a solution that included nuclear staining, DAPI (1:1000), and sealed with nail polish.

For detection of BoNT/A-cleaved SNAP-25 in the frozen human brain (positive control, 2 replicates) and vascular tissue (7 replicates), we co-incubated with antibodies against the adrenergic nerve marker, tyrosine hydroxylase (TH, Fig. S3, S4), using a similar protocol except that primary antibody was to be a chicken anti-TH (1:200) or a rabbit anti-A-cleaved SNAP25 (1:200). This latter polyclonal antibody has been raised against the C-terminal peptide of SNAP-25, sequence KADSNKTRIDEANQ (Fig.2A) of SNAP25 which is exposed after BoNT/A cleavage and is hidden in the intact SNAP25.^36^ Immunoexpression of these targets was quantified and compared in arterial graft subsamples collected before and after BoNT/A treatment. Images were exported as JPEG files and analyzed using 346 ImageJ software.

### Western blotting

Total protein was isolated from the human cortex and descending branch of the lateral circumflex femoral artery (DLCFA) tissue samples using the N-PER Neuronal Protein Extraction Reagent (ThermoFisher 87792) and Cell Lysis Buffer (ThermoFisher FNN0011) respectively, each supplemented with phenylmethylsulfonyl fluoride (ThermoFisher 36978) and Pierce Protease and Phosphatase Inhibitors (ThermoFisher A32959) and homogenized with a Dounce tissue grinder. Proteins were denatured with sodium dodecyl sulfate (SDS) and heat while disulfide bonds were reduced with 2-Mercaptoethanol before electrophoretic separation of 20 µg of protein on pre-cast 10%-14% polyacrylamide gels with Tris-Glycine buffer supplemented with Bolt Antioxidant (ThermoFisher BT0005).

Separated proteins were blotted onto activated 0.45 µm PVDF membranes in a Bis-Tris transfer buffer solution with Bolt antioxidant and 10% methanol. Membranes were blocked for 2 hours in Starting Block T20 Tris-buffered Saline (TBS) Blocking Buffer (ThermoFisher 37543) at 4°C with gentle agitation before overnight incubation with primary antibody diluted in Blocking Buffer at 4°C with gentle agitation. Five purified polyclonal rabbit antibodies were incubated at the upper end of the manufacturer-recommended concentration with corresponding dilutions as follows: rabbit anti-ROCK-1/2 (Millipore Sigma 07-1458) at 1:500 dilution, rabbit anti-Myosin Light Chain phospho-specific (Ser19) (Millipore Sigma AB3381) at 1:1000 dilution, rabbit anti-phospho-MYPT1 (Thr850) (Millipore Sigma 36-003) at 1:500 dilution, rabbit anti-GAPDH (Millipore Sigma G9545) at 1:5000 dilution, and rabbit anti-SNAP25 (Millipore Sigma S9684) at 1:5000 dilution. The non-commercial rabbit anti-cleaved-SNAP25 antibody was incubated at 1:500 and 1:1000 dilution. Blots were probed with goat anti-rabbit IgG (H+L)-HRP conjugate (Bio-Rad 1706515) at 1:3000 dilution for 1 hour after three 10-minute washes in TBS with 0.05% Tween 20 detergent. After six more 5-minute washes blots were developed with Pierce Chemiluminescent substrate solution (Thermo Fisher 34580) and imaged. Images were exported as JPEG files and analyzed using 346 ImageJ software.

### Assessing BoNT/A effects on selected neurotransmitter (NT) levels

Liquid chromatography-mass spectrometry (LC-MS) was used to determine whether BoNT/A administration altered NT levels in the arterial graft wall before and after BoNT/A exposure. Adventitia were removed from the specimens that were used for NT level measurements to exclude those NT which were released from the nerve terminals. Briefly, the stock solutions of individual standards were made by weighing approximately 1mg of the original powder for metanephrine, norepinephrine, epinephrine, and normetanephrine standards (bought from Sigma-Aldrich (St. Louis, MO)) and dissolving in 50% methanol to achieve a concentration level of 1.0 mg/mL. The stock solution was prepared by mixing the individual standards to make a working solution of 50 ug/mL each. The working solution was used for calibration curves and quality control standards.

The tissue samples were thawed on ice, ∼20-60 mg of thawed tissue was sliced off and placed in a 2 mL screw cap microcentrifuge tube (VWR, Radnor, PA) containing a 5 mm i.d. clean stainless-steel beads (Qiagen, Hilden, Germany). To every 20-60 mg of tissue, 500 μL of ice-cold MeOH, acetonitrile, and Water (2:2:1) was added. To this suspension, 10 μL of Voriconazole (internal standard at 50 ng/mL) was added. The samples were homogenized on a TissueLyzer (Qiagen, Hilden, Germany) at 50 Hz for 5 min and centrifuged at 12 000 rpm for 10 min. About 400 uL of the supernatant of the tissue extracts were transferred into another microcentrifuge tube and then evaporated to dryness under a steady stream of filtered and dried N2 gas. The dried residues were then reconstituted with 50% methanol (50 μL) and centrifuged to remove any undissolved debris. The analytes were separated and quantified with Agilent 1290 UPLC linked onto a QTRAP Sciex 6500+ LCMS/MS system. For separation, a Kinetex F5 column (2.6 μm, 2.1 ×150 mm, Phenomenex) was used with the following binary mobile phase: water + 0.01% FA (mobile phase A) and methanol + 0.01% FA (mobile phase B). The gradient [time (%A/%B)] was programmed as follows: 0 min (99.8/0.2)–2 min (99.8/0.2)–5 min (98/2)– 11 min (75/25)–13 min (2/98)–17 min (2/98)–18 min (0.2/99.8)–20 min (0.2/99.8). An injection volume of 10 μL was used, and the flow rate was held at 0.2 mL/min while the column temperature was maintained at 30°C. Each of the targeted analytes was identified and quantified using their unique MRM signatures (m/z 180.1/148.1 for metanephrine; 152.1/107.1 for norepinephrine; 166.1/107.1 for epinephrine and 166.1/106 for normetanephrine) in positive mode (ESI). The analyses were conducted in electrospray ionization (ESI) positive mode with curtain gas flow rate set as 35 L/min, Gas 1 flow rate at 60 L/min; Gas 2 gas flow rate at 70 L/min, spray voltage at 2.5 kV, and source temperature at 500 °C.

### Statistical analysis

Description and statistical analysis were performed using JMP Pro 15 (SAS Institute Inc., Cary, NC, USA) and R 4.3.1 (R Foundation for Statistical Computing, Vienna, Austria). For description, continuous variables are described using mean and standard deviation (SD) or median and interquartile range (IQR), with binary and categorical variables described using counts and percentages. The kappa method was used for the assessment of inter-observer reliability. The two groups were compared using generalized linear models for continuous variables and logistic regression or multinomial logistic regression for binary or categorical variables, respectively. Analysis of measurements comparing BoNT/A treated and untreated samples from the same subjects were done with generalized linear mixed-level models to account for expected intra-subject autocorrelation. Families and links were selected by comparison by the Akaike Information Criterion (AIC).^37^ The R packages nnet,^38^ glmmTMB,^39^ and MuMIn^40^ were used for multinomial and mixed-level modeling, respectively. Changes in mRS from baseline to follow-up were compared using a mixed-level generalized linear model, clustered by patient.

## Data Availability

All data produced in the present study are available upon reasonable request to the authors
All data produced in the present work are contained in the manuscript

## Non-standard Abbreviations and Acronyms

5HIAA: 5-hydroxyindoleacetic acid
BoNT: botulinum toxin
DLCFA: descending branch of the lateral circumflex femoral artery
EC-IC: extracranial-to-intracranialI
glmm: generalized linear mixed-level models
IC-IC: intracranial-to-intracranial
IF: immunofluorescence
IHC: immunohistochemistry
IQR: interquartile range
LC-MS: liquid chomotography-mass spectrometry
MAP: mean arterial pressure
mRS: modified Rankin Scale
NT: neurotransmitter
pMLC: phosphorylated myosin light chain
pMYPT1: phosphorylated myosin phosphatase target subunit 1
RAFCF: radial artery fasciocutaneous flow-through free flap
RAFF: radial artery fascial flow-through free flap
RAG: radial artery
rho kinase: ROCK1/2
SAH: subarachnoid hemorrhage
SD: standard deviation
SDS: sodium dodecyl sulfate
SNAP25: synaptosomal-associated protein 25 kDa
SRM: selected reaction monitoring
TBS: Tris-buffered saline
TH: tyrosine hydroxylase
VMA: vanillylmandelic acid
VSMCs: vascular smooth muscle cells.: 

## Acknowledgment

All original artwork for this manuscript was created by Studio Kayama LLC.

## Author contributions

Conceptualization: NAA, KR, SS, RR, JC, JJR

Methodology: NAA, KR, RR, AA, GTK, SL, IA, OR, JJR

Investigation: NAA, KR, AA, SS, GTK, FF, LS, IA, DH, OR, JC, JJR

Visualization: NAA, KR, AA, SS, GTK, FF, VN, DH, OR, JJR

Funding acquisition: NAA, KR, RR, JC, JJR

Project administration: NAA, KR, SS, LS, OR, JJR

Supervision: NAA, SS, RR, SL, LS, DH, OR, JC, JJR

Writing – original draft: NAA, KR, SS, RR, AA, JJR

Writing – review & editing: NAA, KR, SS, RR, GTK, FF, VN, SL, IA, DH, OR, JC, JJR

