## Supplemental Figures and Tables for "Botulinum Toxin Application for Treatment of Graft Vasospasm: A Reverse Translational Study"

**Fig. S1. Identifying characteristics of vasospasm.** Radiographic images of Cerebral circulation illustrating A) Non-spasm and B) Spasm vessels. Spasm is indicated by constricted diameter (white arrow).

**Fig. S2. Full-length SNAP25 immunohistochemistry and brain H&E Staining.** IHC of Cortex (panels I, III) and Artery (Panels II and IV). Full-length SNAP25 levels are low in the arteries (1:200) compared to the brain (dilution 1:5000).

**Fig. S3. TH Ab optimization.** Neuroendocrine chromaffin cells, responsible for the biosynthesis of catecholamines, are located throughout the brain (adrenergic neurons) and in the adrenal glands (chromatin cells). The highest density of chromaffin cells is located within the adrenal medulla, the most functionally significant area of catecholamine production.^64^ TH staining (TH, green, and DAPI blue). is absent in the negative Liver samples. Immunofluorescence of representative human liver and adrenal gland at various magnifications (20x and 40x).

**Fig. S4. Immunofluorescence of representative human brain cortex sample with BoNT/A treatment.** Full-length SNAP24 (Fig. S2) is highly expressed in cortical adrenergic neurons (TH, green). Samples treated with BoNT/A result in the production of cleaved SNAP25 (cSNAP25, red). Double staining for TH and cSNAP25 identifies the colocalization of cSNAP25 with TH in cortical adrenergic neurons (Merged, yellow).

**Fig. S5. Cleavage of mouse and human SNAP25 in extracts from the cerebral cortex by BoNT/A.** Extracts were prepared, treated with BoNT/A, subject to SDS-PAGE, and transferred to PVDF membrane, then probed with primary and secondary antibodies as described in the main text. A) Constant (10 Units/mL) BoNT/A applied to different amounts of human cortical proteins. B) Different amounts of BoNT/A applied to a constant amount of human cortical proteins.

**Fig. S6. Brain and arterial morphology microphotographs.** H&E staining of sample human tissue was carefully selected based on intact cortical and vascular morphology and vessel size to reliably detect cSNAP25 and TH before and after treatment with BoNT/A. Note that, the same artery but a different segment was used for treatment.

Fig. S1

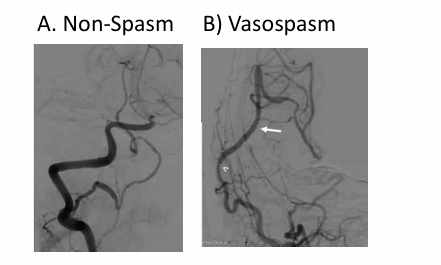

Fig. S2

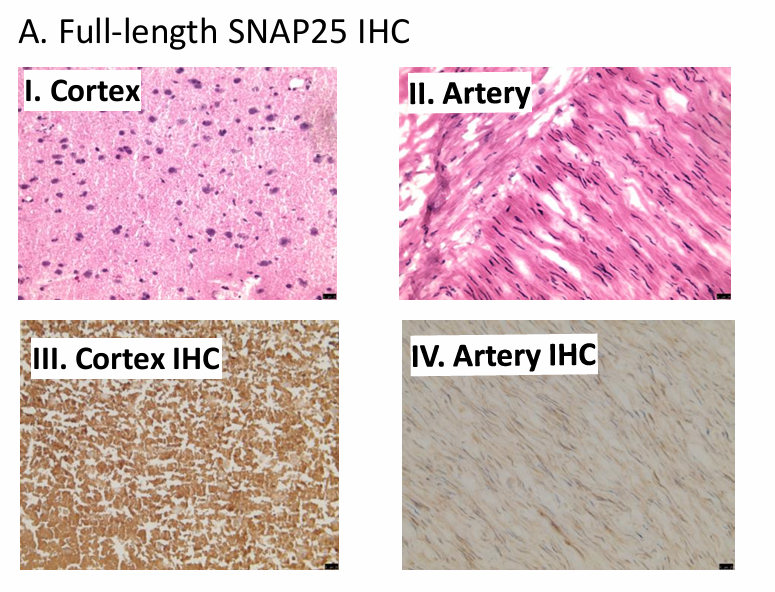

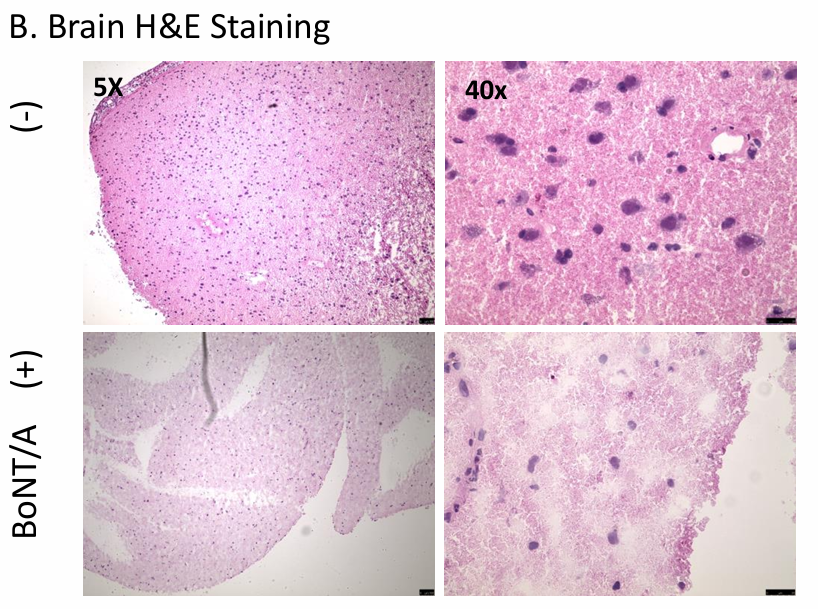

Fig. S3

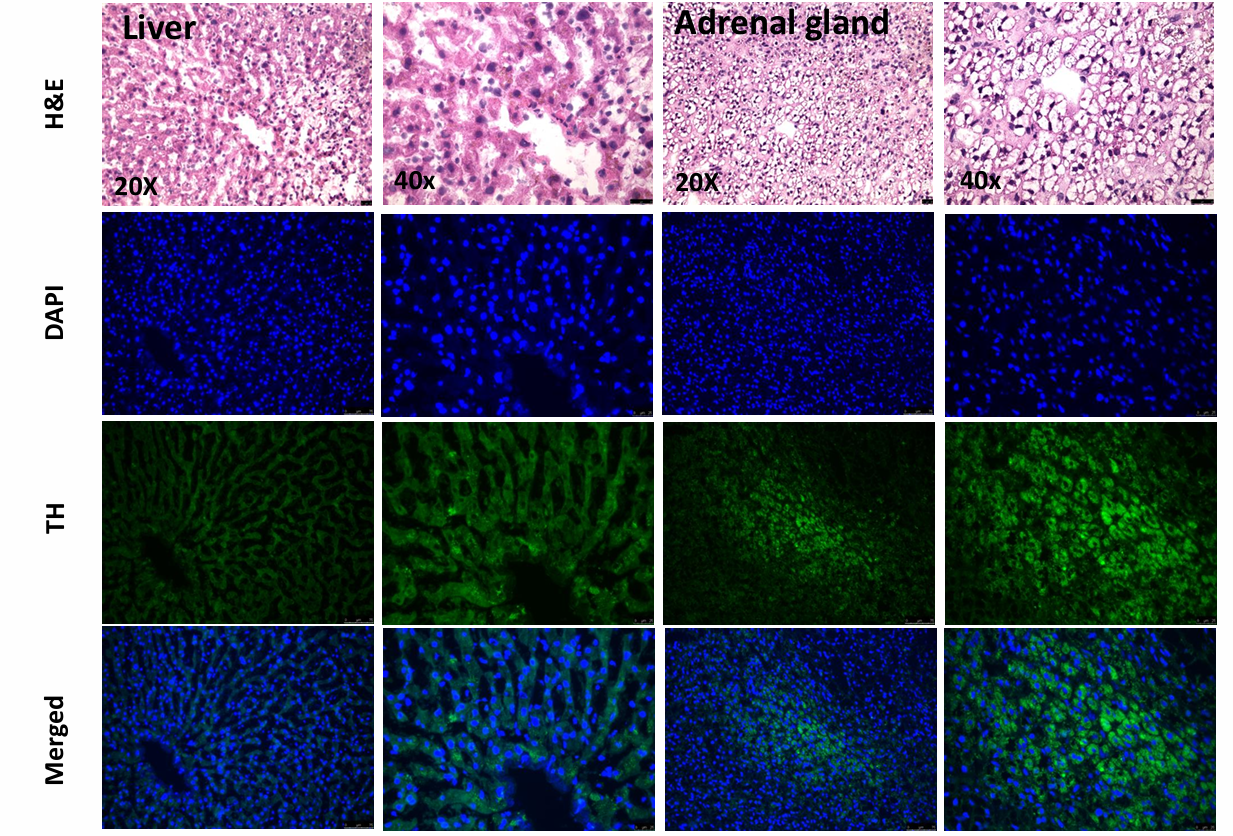

Fig. S4

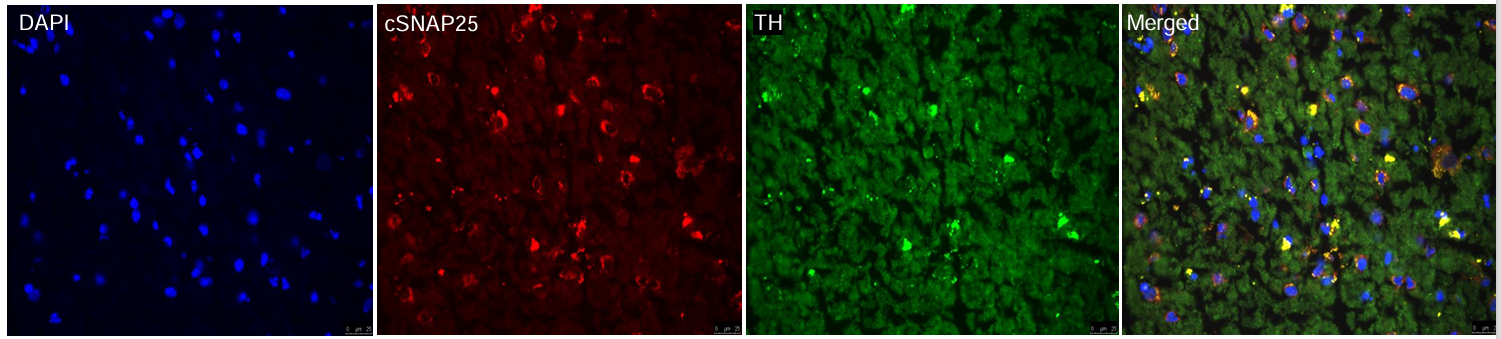

Fig. S5

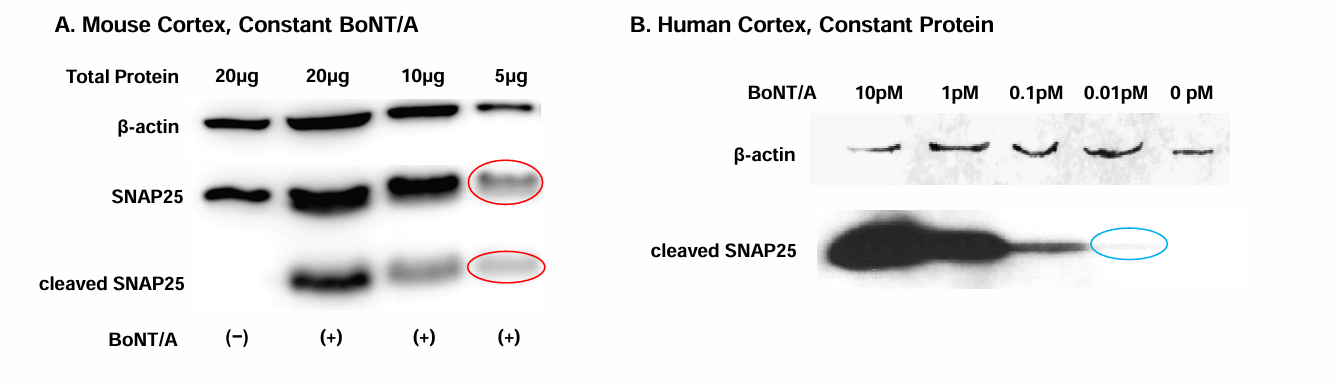

Fig. S6

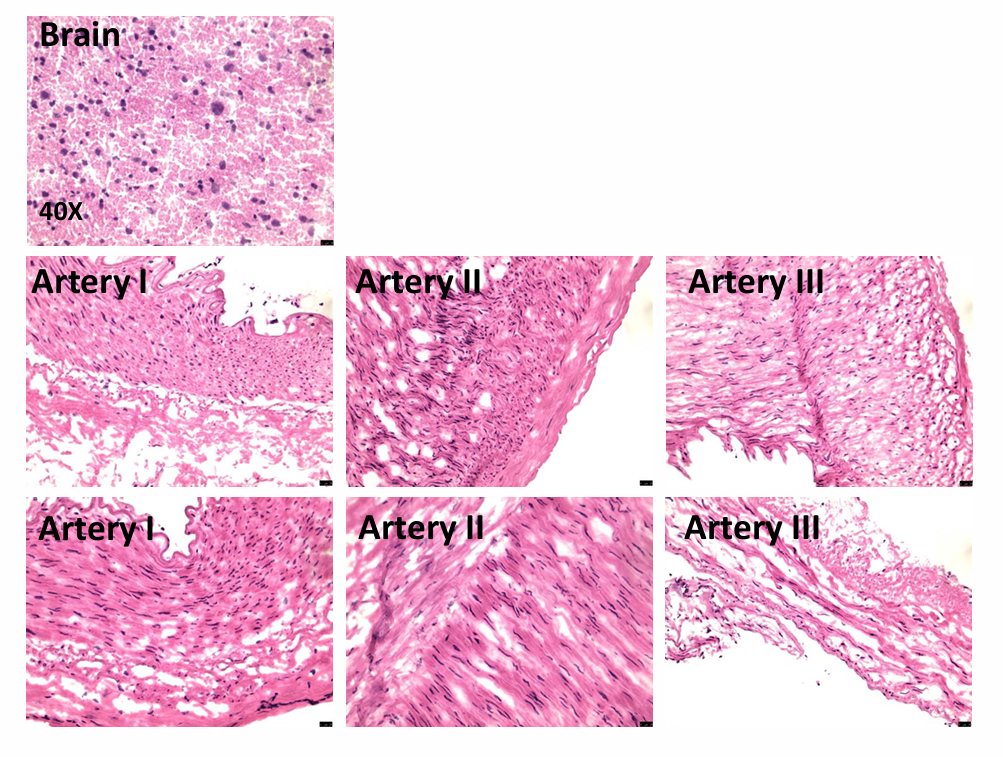

**Effect of varying normalizing protein on western blots**

We recognize western blotting of ROCK pathway proteins used two normalizing standards, specifically β-actin and GAPDH. While both are "housekeeping" proteins, each takes part in distinct biological activities and cannot be trivially presumed to be interchangeable. However, the small number of samples (n = 6 BoNT/A−, 6 BoNT/A+) means that explicitly testing effects of variables beyond BoNT/A treatment is not reliable, as this violates the "one-in-ten rule",^37^ which is a commonly used rule of thumb that models of continuous data can support no more than one parameter for every ten data points. However, this is a rule of thumb and need not be applied inflexibly. Therefore, to account for possible effects of differing normalizing proteins, we examined alternate models that included β-actin/GAPDH as an additional factor, along with possible interaction with BoNT/A status.

We compared candidate models with second-order Akaike information criterion (AICc) for pMLC, pMYPT1, and ROCK1/2. We found that the most parsimonious model for pMLC was the model featured in the main text. The most parsimonious model that included normalizing protein was "pMLC ~ BoNT/A + normalizer + (1 | Subject)" (Table S1). This model still predicted a significant role for BoNT/A vs. pMLC levels, but normalizing protein also had a significant effect. The probability of this model being "true" vs. the BoNT/A only model was 21.5%, based on differences in AICc.^38^ The most parsimonious model for pMYPT1 was also the model used in the main text. The most parsimonious model that included normalizing protein was "pMYPT ~ BoNT/A + normalizer + (1 | Subject)". This model did not estimate a significant effect for the normalizing protein difference. Estimated probability of this model from AICc was 6.3%. By contrast, the most parsimonious model for ROCk1/2 included the normalizing protein difference and was "ROCK1/2 ~ BoNT/A + normalizer + (1 | Subject)". In this model, the normalizer had a significant effect, but BoNT/A still had no significant effect (Table S1). In addition, the probability for this model vs. the model in the main text was 95.8%. We acknowledge that using different western blot normalizing proteins for a single experiment is highly problematic. However, we also have some confidence in our results because taking this event into account did not alter the significance (or lack) of BoNT/A treatment.

**Table S1. Models that include normalizing protein difference.**

| **Response** | **Effect** | **χ^2^ (df)** | **p value** |
| --- | --- | --- | --- |
| pMLC | BoNT/A  normalizer | 53.847 (1)  5.192 (1) | **< 0.001**  **0.023** |
| pMYPT1 | BoNT/A  normalizer | 26.685 (1)  0.951 (1) | **< 0.001**  0.330 |
| ROCK1/2 | BoNT/A  normalizer | 3.366 (1)  53.436 (1) | 0.067  **< 0.001** |
